# Diagnostic efficacy of leukocyte esterase dipstick in diagnosing spontaneous bacterial peritonitis among cirrhotic patients in tertiary hospitals, Dodoma, Tanzania

**DOI:** 10.1101/2023.08.04.23293652

**Authors:** Salum Ali Mwinyi, Emmanuel Sindato

## Abstract

**Background:** Spontaneous bacterial peritonitis (SBP) is complications of end stage liver disease, it associated with morbidity and mortality, the gold standard for diagnosing SBP is an ascitic fluid Polymorphonuclear neutrophil count (PMN) of ≥ 250 cells/mm^3^, this examination is time consuming and costly. Urine reagent dipstick detecting leukocyte esterase activity has been suggested as quick and affordable substitute. The purpose of this study was to evaluate the CYBOW™ 10 strip’s ability to diagnose SBP.

**Methods:** A Crossectional analytical study was conducted from November 2022 to June 2023. 224 patients with confirmed cirrhotic ascites, aged ≥ 18 years and met inclusion criteria were recruited in the study. By following sterile procedure ascitic fluid was collected, bedside ascitic fluid examination by CYBOW™ 10 reagent strips, and the samples for cytological examination were taken. Ascitic fluid with PMN ≥ 250cells/mm^3^ was considered positive for SBP, and +1 to +3 of CYBOW™ 10 reagent strip, was used as cut off levels for a positive SBP. By using SPSS version 25, 2 by 2 table was applied to determine the sensitivity (Sn), Specificity (Sp), Positive Predictive value (PPV), and Negative Predictive Value (NPV), and Receiver Operating Characteristic (ROC) was applied to determine the area under the curve of the leukocyte esterase dipstick.

**Results:** On the basis of the ascitic fluid PMN count, 42 (18.75%) individuals had SBP. At cut-off point of +2 CYBOW™ 10 urine reagent strip had Sensitivity of 82.14%, Specificity of 99.39%, PPV of 95.83% and NPV of 96.79%, with AUC of 0.9074.

**Conclusion:** CYBOW™ 10 reagent strip test might be a helpful tool for doctors, when a rapid cell count for SBP diagnosis is not available. These strips are readily available and inexpensive, can be very helpful in areas with low resources and to patients who are unable to pay for cytology.

## INTRODUCTION

Spontaneous bacterial peritonitis (SBP) is a bacterial infection of ascitic fluid in the absence of focal source of infection, occurs mainly in individual with cirrhotic liver (1). SBP is one of the severe complication associated with liver cirrhosis that has high impact on quality of life and increase in mortality (2,3). Globally, the prevalence of SBP is about 17.12 % (4), based on individual studies, the prevalence of SBP is ranging from 6.7% to 67.7% (5,6). It was established that after a single episode of SBP, recurrence and mortality rates increases significantly (7), and estimated one year survival of SBP without treatment is ranging from 30 to 50 percent with an estimated two year survival is ranging from 25 to 30 percent (8), however with early treatment the death rate could be as low as 5 percent (9). Diagnosis of SBP is established by cellular counting with Polymorphonuclear cell being equal or above 250 cell/mm^3^ (10,11). This examination is time-consuming and costly. Multiple studies have attempted to find alternative way for diagnosing SBP, like the use Procalcitonin-Erythrocyte Sedimentation Rate-C-reactive protein (PEC) index (12), interferon gamma induced protean, homocystein, procalcitonin and C-reactive protean (13), ascitic calprotectin (14), and leucocyte esterase dipstick(15).

Leukocyte esterase is an enzyme produced by activated neutrophil, usually found in the infected fluids in the body, leukocyte esterase reagents commonly used in the diagnosing UTI (16). Leukocyte esterase dipstick is fixed with 3-Hydroxy-5-phenyl-pyrrole that has been esterified with an amino acid, hydrolysis of this ester by the esterase released by activated neutrophil produce 3-hydroxy-5-phenyl-pyrrole, this reacts with a diazonium salt embedded in the dipstick to produce purple color (17), the intensity of color correlate with number of neutrophil in the fluid (18). different studies have shown the promising results of the efficacy of using dipstick as cheap and rapid alternative test in diagnosing SBP, nonetheless these studies achieved different level of sensitivity, specificity, PPV and NPV, for instance multiple studies have shown good sensitivity and specificity of leucocyte esterase dipstick in diagnosing SBP (19–22), however there are studies argue that leucocyte esterase dipstick is not appropriate alternative of cytology for diagnosing SBP (23,24).

This study evaluated the efficacy leucocyte esterase dipstick in diagnosing SBP.

### Patients and methods

We conducted a cross-sectional analytical study to determine the efficacy of leukocyte esterase dipstick in diagnosing SBP among cirrhotic patients admitted in Tertiary Hospitals, Dodoma, Tanzania. Patients with secondary bacterial peritonitis, patients who had been kept on antibiotics for more than 12 hours at the time of recruitment, patients who had history of abdominal surgery in the last 4 weeks and those with diagnosis of peritoneal malignancy, were excluded from this study. The study included 224 patients with confirmed liver cirrhosis of various etiologies, admitted between November 2022 and June 2023. Upon recruitment they were informed on the nature of the study, procedures and testing to be done and were informed that their participation was optional and that they might opt-out anytime. Privacy and confidentiality were maintained by substituting participants’ names with identification numbers; however, their standard of care was unaffected by their decision to participate. Patients who were diagnosed to have SBP received treatment according to the standard guidelines.

The ascitic fluid was be aspirated in the flanks, a little outside the mid-point of a line drawn from the umbilicus to the anterior superior iliac spine. The skin at the chosen point was cleaned using 70% alcohol then was infiltrated with local anesthetic, and the anesthetic was then be injected down to the parietal peritoneum, and ascitic fluid was withdrawn using an 18-gauge cannula, ten milliliters of the sample was inoculated into a heparin anticoagulant tube which was sent for cytology examination and ten milliliters was used for dipstick esterase testing.

Soon after paracentesis, sample of ascitic fluid was placed in a in a dry, clean container that is normally used to collect urine, CYBOW™ 10 urine reagent strip was used to determine leucocyte esterase activity, the dipstick strip contains four colorimetric level scales (negative, 1+, 2+ and 3+), the strip was taken from the closed canister and was used immediately and the canister was closed tightly instantly. The reagent areas of the strip were immersed in a fresh well-mixed ascitic fluid and were immediately removed to avoid dissolving reagents. The edge of the strip was run against the rim of the container while removing it to remove excess ascitic fluid. The strip was held in a horizontal position and the edge of the strip was brought into contact paper towel to avoid mixing chemicals from adjacent reagent area and/or soiling hands with ascitic fluid. The reagent was then compared to the corresponding color blocks on the canister label after 90 seconds as described by manufacturer, each reading was graded by 2 well trained medical personnel, who were unaware of the results of ascitic cytological examination at the time of reading, and if there was a discrepancy, third trained personnel was called to solve the discrepancy by repeating ascetic fluid dipstick reading and grade the results, the manufacturer indicated the following link between PMN cell count and the 4-grade scale: negative, 0 PMN cell/mm^3^ 1+, ≥25 PMN cells/mm^3^; 2+, ≥ 75 PMN cells/mm^3^; 3+, ≥ 500 PMN cells/mm^3^, when a dipstick recorded +1, +2, +3 was considered to be positive, and the patient was labeled SBP positive. Negative dipstick tests were defined as negative results on the colorimetric scale.

PMN cell count was performed on ascitic fluid as per standard procedure. Samples was collected into a heparin anticoagulant tubes, sample was analyzed by 2 pathologists at the Benjamin Mkapa Hospital (BMH) within four hours after sample extraction, samples that were collected from Dodoma Regional Referral Hospital were transferred to BMH via cool box with ice pack, and were processed within four hour of extraction, in the laboratory the ascitic fluid was be stirred briskly to disperse suspended cells, then the sample was centrifuged for 4 minutes, then 3-4 labeled microscopic glass slides for smearing was used, one to two drops of the sediment was placed on the slide and allow it to spread evenly by placing another slide over it, then gently slides were pulled apart with an easy sliding motion to get alternate thick and thin area, then immediately prepared smears was be submerged in 95 percent ethanol, was fixated for at least 15 minutes, then was stained by using Papanicolaou stain, then all samples were examined by using Leica DM300 microscope equipped with digital camera, ascitic fluid cell count was done by using Chamber Counting Method, with Polymorphonuclear (PMN) cell count of equal or more than 250 cells/mm^3^ was used as a diagnostic cut off level.

The data obtained in this study was be managed and stored confidentially on an excel spread sheet using an encrypted computer. The data was analyzed using SPSS-25, By using 2 by 2 table the sensitivity (Sn), Specificity (Sp), Positive Predictive value (PPV), and Negative Predictive Value (NPV) was determined and to determine the area under the curve of the leukocyte esterase dipstick, Receiver Operating Characteristic (ROC) was applied.

## Ethical Issues

The Vice Chancellor’s office at the University of Dodoma provided permission to conduct the study after obtaining ethical clearance from the Directorate of Research and Publications (reference number MA.84/261/02/10). Following that, the administrative departments of Benjamin Mkapa and Dodoma Regional Referral Hospitals approved data collection with reference numbers AB.150/293/01/196 and MA.84/261/02/’A’/59/8, respectively.

## Results

Two hundred and twenty four participants with a liver cirrhosis and ascites were assessed for presence of Spontaneous Bacterial Peritonitis, 42 (18.75%) patients were found to have SBP by Polymorphonuclear cell count.

Based on the findings of dipstick values, the sensitivity and specificity was determined, the sensitivity of +1 was determined to be 68.75% and specificity was 83.43%, the sensitivity of +2 was determined to be 82.14% and specificity was 83.43%, the sensitivity of +3 was determined to be 37.5% and specificity was 100%. Positive Predictive Value of +1 was determined to be 26.83% and the Negative Predictive Value was found to be 96.79%, for +2 the Positive Predictive Value was found to be 95.83% and Negative Predictive Value was determined to be 96.79%, and for +3 the Positive Predictive Value was determined to 100% and the Negative Predictive Value was found to be 96.79% “Table 1”. According to ROC curve findings, a cut-off of +2 provided the best diagnostic accuracy for the strips “Figure 1”.

**Table 1:** Sensitivity, Specificity, Positive Predictive Value and Negative Predictive Value, CYBOW™ series urine reagent strip for different dipstick values.

|  | +1 | +2 | +3 |
| --- | --- | --- | --- |
| **Sensitivity** | 68.75% | 82.14% | 37.5% |
| **specificity** | 83.43% | 99.39% | 100% |
| **PPV** | 26.83% | 95.83% | 100% |
| **NPV** | 96.79% | 96.79% | 96.79% |

**Figure 1.**
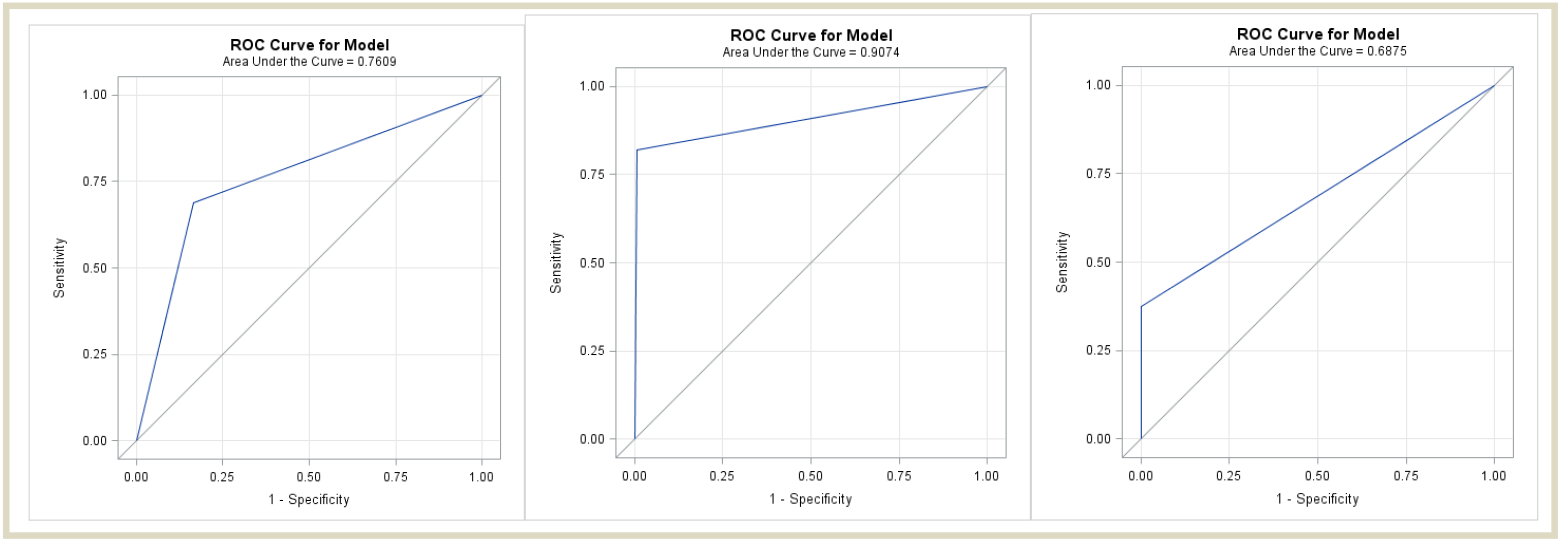
The Receiver Operating Characteristic (ROC) curves at different cut-off levels.

## Discussion

SBP continues to be a significant cause of morbidity and mortality in people with liver cirrhosis (7,25). Earlier diagnosis of SBP and treatment enhance survival in patients with liver cirrhosis, while failure or delay in establishing diagnosis of SBP in patients with liver cirrhosis is associated with increase in mortality (26). Still the Gold standard test for diagnosing SBP is ascitic fluid PMN cell count, with PMN count of equal or more than 250 cells/mm^3^ suggestive of SBP regardless of ascitic culture (27). Because of the cost and limited number of expert, the test is rarely performed in our setting (28,29).

Our study demonstrated the efficacy of Leucocyte esterase dipstick in diagnosing SBP. At +2 cut-off value, it demonstrated the Sensitivity of 82.14%, Specificity of 99.39%, PPV of 95.83% and NPV of 96.79%, with AUC value of 0.9074, For the diagnosis of SBP, performance at +2 cut-off was rather satisfactory, this is comparable to the findings of a study by Chugh et al, was evaluating the efficiency of Multistix 10 SG reagent strip in diagnosing SBP found similar pattern of efficacy with sensitivity of 95%, specificity of 96.4%, PPV of 86.4% and a NPV of 98.8% at a cut-off of grade +2, (21), similarly a study conducted by Bafandeh et al, evaluated the efficacy of Medi-Test Combi strips to diagnose SBP, it had sensitivity of 87.8%, specificity of 96.7%, PPV of 92.3% and NPV of 94.6% for the 2+ cut-off scale, had a comparable results to our strips (30), However there are studies in the literature which gave results different from our findings, for instance a study of Afrida et al, used Combur10 Test®M assessed the efficacy of LER in diagnosing SBP, at a dipstick value of +2, the leukocyte esterase test exhibited comparable specificity of 94% and NPV of 80%, however it showed poor sensitivity of 63.9% with very poor PPV 32% (31), the small number of participants in the study and nature of leucocyte esterase reagent could explain the discrepancy. Moreover a study from France by Nousbaum et al, used Multistix 8SG urine test to evaluate the efficacy of leucocyte esterase dipstick in diagnosing SBP, at dipstick value of +2 found the poor sensitivity of 45.3% and weak PPV of 77.9%, however had excellent specificity of 99.2% and NPV of 96.9% (23). Furthermore there are reported leucocyte esterase reagents in the literature with better diagnostic efficacy compared to reagent strip used in our study, for instance, in the study conducted by Torun et al, which used Aution Sticks 10 EA to assess the efficacy of the LER in diagnosing SBP, at 2+ cut-off point it was found to have better efficacy compared to reagent used in our study sensitivity (93%), specificity (100%), PPV (100%), and NPV (98%) (32), the discrepancy could be explained by the different nature of strips used in the studies as was described by Koulaouzidis in meta-analysis (33).

## Conclusion

Our findings align with previous reports, providing further evidence that the leukocyte esterase reagent strip test is a valuable and efficient tool for the rapid screening of spontaneous bacterial peritonitis (SBP). These strips are readily available and inexpensive, can be very helpful in areas with low resources and to patients who are unable to pay for cytology.

## Data Availability

All relevant data are within the manuscript and its Supporting Information files.

## *A*cknowledgements

I am profoundly thankful to Dodoma regional referral hospital (DRRH) and Benjamin Mkapa Hospital (BMH) for their support, and providing a good and suitable environment for my training, which made this study possible.

## Author Contributions

Conceptualization: Salum A. Mwinyi, Emmanuel Sindato

Data curation: Salum A. Mwinyi.

Formal analysis: Salum A. Mwinyi.

Investigation: Salum A. Mwinyi.

Methodology: Salum A. Mwinyi, Emmanuel Sindato.

Supervision: Emmanuel Sindato.

Writing – original draft: Salum A. Mwinyi.

Writing – review & editing: Emmanuel Sindato.

